# Addressing Socioeconomic Inequities in Children’s Cardiovascular Health via Positive Experiences

**DOI:** 10.1101/2025.07.15.25331608

**Authors:** Shuaijun Guo, Rushani Wijesuriya, David Burgner, Meredith O’Connor, Sharon Goldfeld, Richard S Liu, Naomi Priest

**Affiliations:** Centre for Community Child Health, Murdoch Children’s Research Institute, Melbourne, Australia; Department of Pediatrics, University of Melbourne, Melbourne, Australia; Clinical Epidemiology and Biostatistics Unit, Murdoch Children’s Research Institute, Royal Children’s Hospital, Melbourne, Australia; Inflammatory Origins Group, Murdoch Children’s Research Institute, Royal Children’s Hospital, Melbourne, Australia; Department of General Medicine, Royal Children’s Hospital, Melbourne, Australia; Department of Pediatrics, Monash University, Melbourne, Australia; Melbourne Children’s LifeCourse Initiative, Murdoch Children’s Research Institute, Melbourne, Australia; Faculty of Education, University of Melbourne, Melbourne, Australia; Institute of Endocrinology and Diabetes, Children’s Hospital at Westmead, Sydney, Australia; School of Pediatrics and Child Health, University of New South Wales, Sydney, Australia; The Centre for Social Policy Research, Australian National University, Canberra, Australia; Indigenous Health Equity Unit, Melbourne School of Population and Global Health, University of Melbourne, Melbourne, Australia

**Keywords:** maternal education, positive experiences, health inequities, cardiovascular health, longitudinal, children, interventional effects

## Abstract

**Objectives:** Socioeconomic disadvantage leads to poor cardiovascular health and this relationship may be mediated by positive childhood experiences. This study aimed to estimate the extent to which promoting positive experiences could reduce socioeconomic inequities in children’s cardiovascular health.

**Methods:** Data source: The Longitudinal Study of Australian Children Child Health CheckPoint (N=1874). Exposure: Maternal education (low/medium/high) as a key indicator of family socioeconomic position during pregnancy. Outcome: Cardiovascular health (11-12 years) (poor/good) quantified by four health behaviors and four health factors. Mediator: Multiple positive experiences (≥2/<2) indicated by positive parenting, supportive relationships, environments, and high social engagement (2-11 years). We conducted a causal mediation analysis using an interventional effects approach, adjusting for childhood adversity and other potential confounders.

**Results:** Children with low (risk difference=4.9%, 95% CI=-3.2%, 13.0%) or medium (risk difference=5.6%, 95% CI=-1.2%, 12.5%) maternal education had a higher risk of poor cardiovascular health compared to those with high maternal education. Causal mediation analysis estimated that increasing the levels of positive experiences in children with low or medium maternal education to be like their high maternal education peers could reduce these risk differences by 1.0% (95% CI= −0.8%,1.5%) and 0.5% (95% CI=-0.5%, 1.5%) respectively, reducing cardiovascular inequities by 20.4% and 8.9%.

**Conclusions:** Targeted policy interventions that promote positive experiences are potential opportunities to reduce socioeconomic inequities in children’s cardiovascular health. However, such interventions should be considered within a broader and multipronged approach that includes addressing socioeconomic disadvantage itself and other socially distributed drivers of cardiovascular diseases to achieve the maximum impact.

**Article Summary:** We explore the potential of positive childhood experience interventions to reduce socioeconomic inequities in children’s cardiovascular health.

**What’s Known on This Subject:** Socioeconomic disadvantage is associated with poor cardiovascular health. Positive childhood experiences are emerging as protective factors, but their potential to reduce socioeconomic inequities in children’s cardiovascular health remains unexplored.

**What This Study Adds:** Promoting positive experiences partially reduces socioeconomic inequities in children’s cardiovascular health. An integrated and multi-faceted approach that tackles the diverse drivers of cardiovascular health is essential to achieve the maximum impact.

## INTRODUCTION

Cardiovascular disease (CVD) is the leading cause of mortality globally, accounting for 32% of all deaths in 2020.^1^ The economic burden of CVD is substantial, with global costs estimated at US$1 trillion in 2030.^2^ Socioeconomic disadvantage is a well-established determinant of CVD, contributing to differences in the incidence and mortality of CVD.^3^ Addressing socioeconomic inequities in CVD is a priority of governments worldwide. Evidence suggests that more than 80% of CVD can be prevented or modified in early life by following healthy lifestyles and addressing risk factors such as high blood pressure and diabetes.^4^

The American Heart Association (AHA) introduced the concept of ideal cardiovascular health (CVH) in 2010,^5^ which refers to not merely the absence of CVD but the presence of favorable health behaviors (e.g., no smoking, healthy diet, regular physical activity) and health factors (e.g., normal body mass index, healthy blood pressure and lipid levels). These components were updated in 2022 to reflect the Life’s Essential 8 (LE8).^6^ This paradigm represents a shift in cardiovascular research from a deficit-focus approach to a strengths-based approach.^5^ Monitoring CVH at the population level over the life course is essential for identifying CVH disparities and informing targeted interventions.

Children’s CVH is shaped by the social environments where they live and develop across the life span.^6,7^ Socioeconomic inequities in CVH emerge as early as childhood.^7^ Data from the 2013-2018 US National Health and Nutrition Examination Survey indicate that family income is associated with differences in CVH indicators such as nicotine exposure, body mass index, and diet among children aged 2 to 19 years.^8^ These disparities are driven by an unequal distribution of material resources as well as structural barriers that disproportionately affect children from socioeconomically disadvantaged families.^6,8–10^ Addressing CVH inequities in children is likely to yield greater cost-effective benefits than interventions later in life, given the cumulative impact of early life exposures on long-term health outcomes.^8,11^

The mechanisms linking socioeconomic disadvantage to CVH are complex,^12^ including both adverse and positive experiences. While childhood adversity (e.g., family violence, child abuse) has been well-established as a risk factor of CVH,^13,14^ positive childhood experiences warrant specific focus because they are valued by families and communities, and efforts to promote positive experiences are considered highly acceptable, avoiding stigma and aligning with strengths-based practices and policies.^15,16^ Positive experiences refer to a range of events, activities, or situations that foster flourishing and better health outcomes.^17^ Although variably defined, emerging evidence suggests that positive experiences are associated with better CVH,^14,16,18–20^ with possible pathways such as enhanced self-esteem and lower rates of substance use.^21^

Compared to CVH in adulthood, very few studies have explored CVH in childhood from a life course perspective.^8,22^ While there is increasing evidence showing the benefits of positive experiences, the extent to which promoting positive experiences would reduce socioeconomic inequities in CVH remains unknown. To inform intervention opportunities and policy actions on CVH improvement at the population level, we estimated the extent to which promoting positive experiences could reduce socioeconomic inequities in children’s CVH.

## METHODS

### Data source

We drew on a subset of data from the birth cohort (B-cohort) of the Longitudinal Study of Australian Children (LSAC), which commenced in 2004 when children were aged 0-1 year (n=5107). A two-stage clustered design was employed to select a sample that was broadly representative of the Australian child population except those living in remote areas.^23^ Children were followed up every two years. We drew data when children were aged 0-1 years (Wave 1; n=5107), 2-3 years (Wave 2; n=4606), 4-5 years (Wave 3; n=4386), 6-7 years (Wave 4; n=4242), 8-9 years (Wave 5; n=4085), 10-11 years (Wave 6; n=3764) and 11-12 years (CheckPoint wave; n=1874). The CheckPoint wave was a one-off national-wide cross-sectional physical health and biomarker module, nested between LSAC Waves 6 and 7.^24^ Multiple information sources were utilized, including parent interviews, parent-report and child-report questionnaires.

Despite the requirement for children to attend multi-hour, in-person clinic assessments in the CheckPoint wave, over 1,800 families participated, demonstrating a strong commitment and willingness to invest time and travel resources. We found that children who had lower maternal education, came from Aboriginal or ethnic minority backgrounds, and lived in low socioeconomic status neighborhoods were likely to be missed out (see Supplementary file 1).

### Measures

Our conceptual model (Figure 1) shows the hypothesized causal pathway from maternal education (during pregnancy) to children’s CVH (11-12 years), via positive experiences (2-11 years) as an intervention target of interest, informed by current knowledge (see Supplementary file 2). Figure 1 was used to guide the selection of measures and inform the analytic approach.

**Figure 1.**
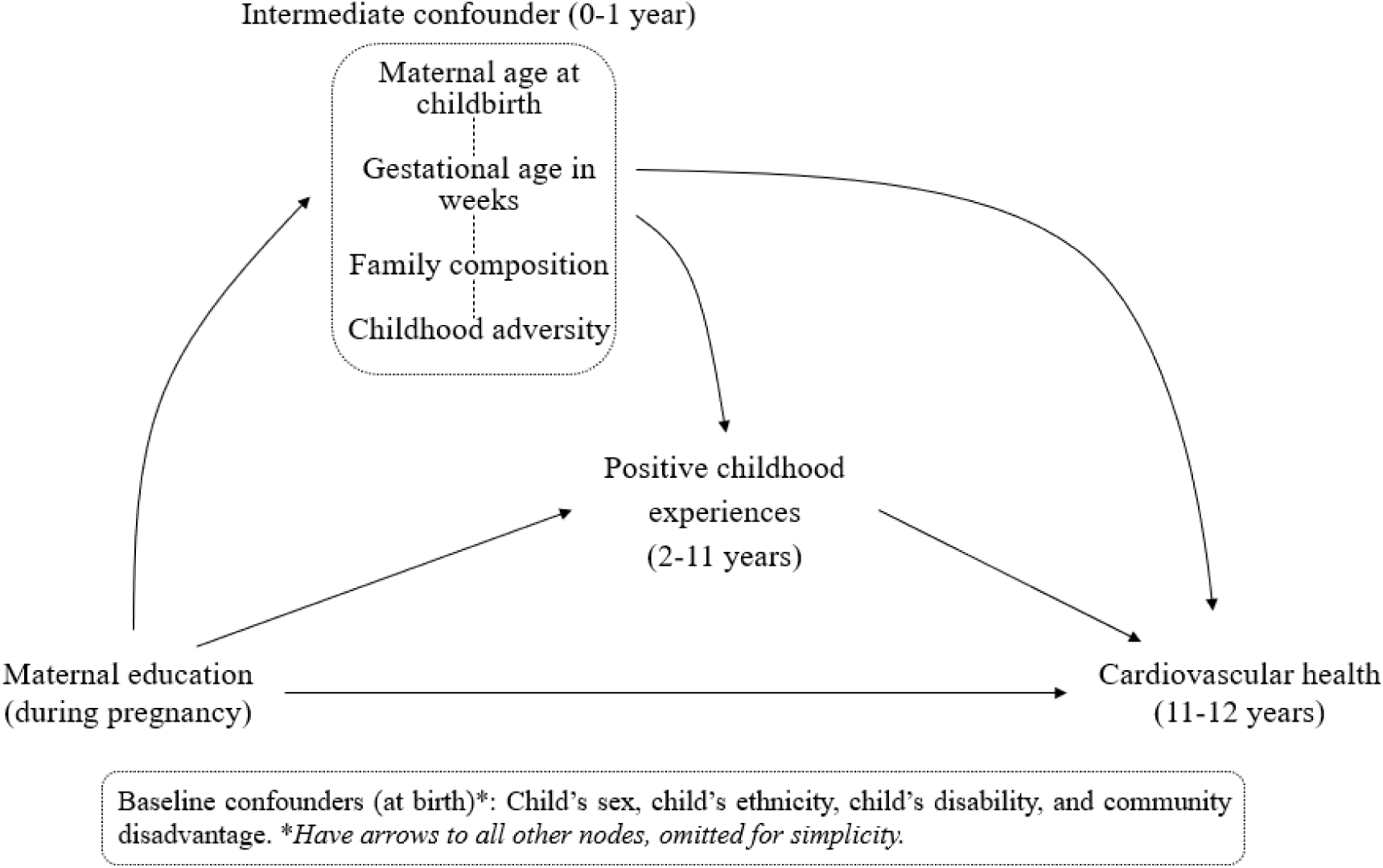
Directed acyclic graph depicting the assumed causal model conceptualizing the pathway from maternal education to children’s cardiovascular health via positive childhood experiences.

#### Exposure (during pregnancy)

Maternal education at Wave 1 was used as a key indicator of socioeconomic resources in the family environment during pregnancy, assuming maternal education did not change significantly from pregnancy to just after birth.^25^ In keeping with previous studies,^25^ we categorized it into three groups: low (Year 12 or below); medium (Certificate I/II/III/IV or Advanced Diploma); and high (Bachelor’s degree or above).

#### Mediator (2-11 years)

Informed by the Health Outcomes from Positive Experiences (HOPE) framework^26^ and previous validation work,^17^ we quantified overall positive experiences using 17 indicators, each mapping to one of the four domains of positive experiences prospectively collected from 2 to 11 years: (1) positive parenting practice; (2) trusting and supportive relationships; (3) supportive neighborhood and home learning environments; and (4) social engagement and enjoyment (see Supplementary file 3). Each indicator was first dichotomized (yes/no) using the top quartile to indicate the presence of exposure to a positive experience at each wave.^17^ Next, we summed the number of positive experiences at each wave for each domain and dichotomized this count at ‘two or more’ to indicate multiple positive experiences in each domain at each wave. To measure multiple positive experiences across four domains at each wave, we summed the number of positive experiences at each wave and dichotomized this count at ‘two or more’ at each wave. Finally, to measure multiple positive experiences over the follow-up period (2-11 years), we summed the number of positive experiences across these waves and dichotomized this count at ‘two or more’, given that a cluster of positive experiences is likely to have a cumulative benefit on health.^27^

#### Outcome (11-12 years)

We quantified CVH at 11-12 years using the LE8 metrics, including four health behaviors (diet, physical activity, cigarette smoking, and sleep) and four health factors (body mass index, non-high-density lipoprotein, blood pressure, and blood glucose). For each child, each of the LE8 metrics was scored on a scale of 0 to 100 (see Supplementary file 4). We then calculated an overall CVH score, using the average value across all eight metrics. According to the AHA recommendation,^6^ we dichotomized the overall CVH score using “0 to 79” to indicate children with poor CVH.

#### Confounders

##### Baseline confounders (at birth)

We posited four baseline confounders: child’s sex (male/female), child’s ethnicity (Anglo or European/minoritized ethnic group/Indigenous), child’s disability status (yes/no), and neighborhood socioeconomic status (top 75% - not disadvantaged/bottom 25% - disadvantaged) assessed by the Socioeconomic Indexes for Areas of Relative Socioeconomic Advantage and Disadvantage.^28^ Due to the possible overlap between maternal education and neighbourhood socioeconomic status, we conducted sensitivity analyses by removing neighborhood socioeconomic status as a baseline confounder.

##### Intermediate confounder (0-1 year)

We posited four intermediate confounders: gestational age in weeks (<37 weeks/≥37 weeks), maternal age at childbirth (<27 years/≥ 27 years), family composition (single parent/two parents), and multiple childhood adversities (<2/≥2) measured by parent legal problems, family violence, household member mental illness, household member substance abuse, harsh parenting, parental separation, unsafe neighborhood, and family member death (see Supplementary file 3).^27^

### Statistical analysis

The analytic sample consisted of all children who attended CheckPoint (N=1874). Participant characteristics were summarized overall and by maternal education, using descriptive statistics. Preliminary analyses were first conducted to confirm whether data were consistent with the expected associations depicted in Figure 1. Specifically, generalized linear models with a log-Poisson link were used to examine unadjusted and confounder-adjusted associations between exposure, mediator and outcome. Descriptive analyses and preliminary analyses were conducted using Stata 18.0.

In all analyses, we ignored the clustering due to postcodes as the correlations between the outcome measures within the postcodes were negligible (intra-cluster correlation= 0.0005). We also did not incorporate the CheckPoint sampling weights in the analyses,^29^ as the incorporation of sampling weights appropriately in the causal mediation approach used below is still an ongoing area of research.

#### Causal mediation analysis

We then conducted a casual mediation analysis using an interventional effects approach to answer the causal question of interest:^30,31^ *what would be the reduction in risk of poor CVH if we could offer effective interventions that promote positive childhood experiences among children with low or medium maternal education?* As well-defined interventions that can collectively address the composite measure of positive experiences are not available in the community, we examined the question by conceptualizing hypothetical interventions that map to a ‘target trial’.^32^

We first estimated the confounder-adjusted absolute difference in the risk of poor CVH in children with low or medium maternal education compared to their high maternal education peers, separately, using *g*-computation.^30,31^ These adjusted differences provided estimates of the overall CVH inequities that we sought to reduce.

Next we evaluated the reduction in risk of poor CVH that would be achieved by a hypothetical intervention that would shift the distribution of the positive experiences in children with low or medium maternal education, to be similar to that in children with high maternal education, using an extended g-computation estimation procedure (see Supplementary file 5).^32^ This provided estimates of the absolute risk differences achieved if we could offer an intervention that promotes positive experiences among children with low or medium maternal education.

The difference between the initial overall CVH inequities and the reduction in risk achieved by the hypothetical intervention provides an estimate of CVH inequities that would remain after the hypothetical intervention. We also report the relative reductions in the gap achieved for children with low or moderate maternal education using these estimates (i.e. reduction in risk of poor CVH achieved by the hypothetical intervention as a ratio of existing socioeconomic inequities in CVH). Standard error estimates were computed using a bootstrap procedure. All mediation analyses were implemented using R Statistical Software 4.3.1 using R package *medRCT*.^33,34^

#### Missing data

In the analytic sample, the percentage of missing data across any of the study variables was 59%. We used multiple imputation by chained equations to reduce bias due to incomplete records, under the missing at random assumption.^35,36^ Imputations of incomplete variables were carried out at the composite level where applicable rather than at the item level due to convergence not being achieved. The imputation model included all study variables and four auxiliary variables (birth weight, GlycA, family income, and parents’ disability status) as well as all two-way interactions amongst exposure, mediator, outcome, and confounders.^37^ Based on the percentage of missing data,^36^ we produced 60 imputed datasets and used Rubin’s rules to obtain the final imputed estimates of interest.^38^ Results using multiply imputed data are shown for preliminary analyses and causal mediation analyses.

## RESULTS

### Sample characteristics

Participant characteristics are summarized in Table 1. At 11-12 years, around half (53.6%) of children had poor CVH. A larger proportion of children with low or medium maternal education had poor CVH compared with their high maternal education peers (low: 55.0%, medium: 57.4%, high: 50.5%). At 2-11 years, a larger proportion of children with low or medium maternal education had fewer positive experiences (low: 63.4%, medium: 55.3%), compared with those with high maternal education (high: 51.3%).

**Table 1.**
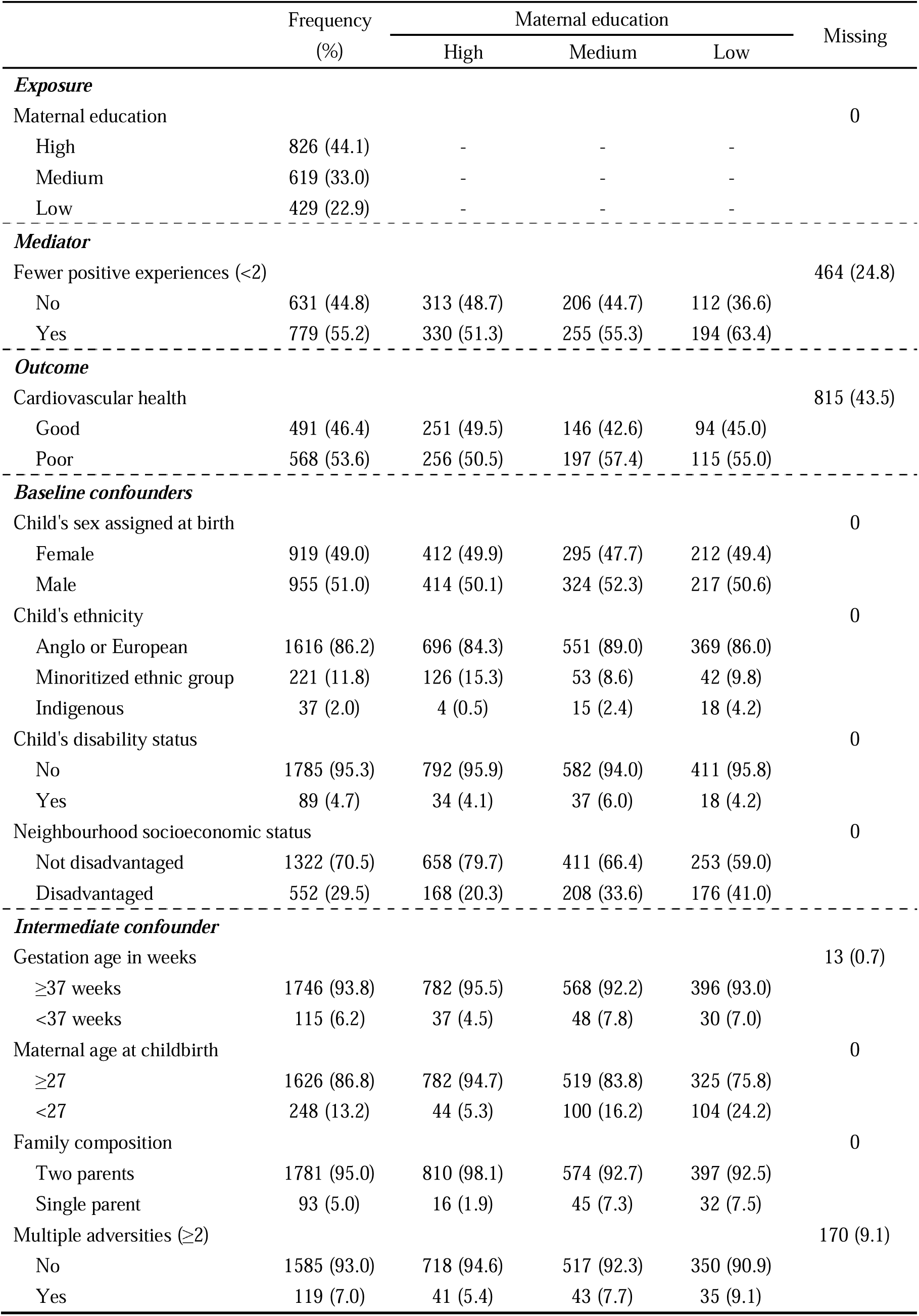
Descriptive information for all study variables in our sample (N=1874). Observed data are shown.

|  | Frequency<br>(%) | Maternal education |  |  | Missing |
| --- | --- | --- | --- | --- | --- |
|  |  | High | Medium | Low |  |
| ***Exposure*** |  |  |  |  |  |
| Maternal education |  |  |  |  | 0 |
| High | 826 (44.1) | - | - | - |  |
| Medium | 619 (33.0) | - | - | - |  |
| Low | 429 (22.9) | - | - | - |  |
| ***Mediator*** |  |  |  |  |  |
| Fewer positive experiences (<2) |  |  |  |  | 464 (24.8) |
| No | 631 (44.8) | 313 (48.7) | 206 (44.7) | 112 (36.6) |  |
| Yes | 779 (55.2) | 330 (51.3) | 255 (55.3) | 194 (63.4) |  |
| ***Outcome*** |  |  |  |  |  |
| Cardiovascular health |  |  |  |  | 815 (43.5) |
| Good | 491 (46.4) | 251 (49.5) | 146 (42.6) | 94 (45.0) |  |
| Poor | 568 (53.6) | 256 (50.5) | 197 (57.4) | 115 (55.0) |  |
| ***Baseline confounders*** |  |  |  |  |  |
| Child's sex assigned at birth |  |  |  |  | 0 |
| Female | 919 (49.0) | 412 (49.9) | 295 (47.7) | 212 (49.4) |  |
| Male | 955 (51.0) | 414 (50.1) | 324 (52.3) | 217 (50.6) |  |
| Child's ethnicity |  |  |  |  | 0 |
| Anglo or European | 1616 (86.2) | 696 (84.3) | 551 (89.0) | 369 (86.0) |  |
| Minoritized ethnic group | 221 (11.8) | 126 (15.3) | 53 (8.6) | 42 (9.8) |  |
| Indigenous | 37 (2.0) | 4 (0.5) | 15 (2.4) | 18 (4.2) |  |
| Child's disability status |  |  |  |  | 0 |
| No | 1785 (95.3) | 792 (95.9) | 582 (94.0) | 411 (95.8) |  |
| Yes | 89 (4.7) | 34 (4.1) | 37 (6.0) | 18 (4.2) |  |
| Neighbourhood socioeconomic status |  |  |  |  | 0 |
| Not disadvantaged | 1322 (70.5) | 658 (79.7) | 411 (66.4) | 253 (59.0) |  |
| Disadvantaged | 552 (29.5) | 168 (20.3) | 208 (33.6) | 176 (41.0) |  |
| ***Intermediate confounder*** |  |  |  |  |  |
| Gestation age in weeks |  |  |  |  | 13 (0.7) |
| ≥37 weeks | 1746 (93.8) | 782 (95.5) | 568 (92.2) | 396 (93.0) |  |
| <37 weeks | 115 (6.2) | 37 (4.5) | 48 (7.8) | 30 (7.0) |  |
| Maternal age at childbirth |  |  |  |  | 0 |
| ≥27 | 1626 (86.8) | 782 (94.7) | 519 (83.8) | 325 (75.8) |  |
| <27 | 248 (13.2) | 44 (5.3) | 100 (16.2) | 104 (24.2) |  |
| Family composition |  |  |  |  | 0 |
| Two parents | 1781 (95.0) | 810 (98.1) | 574 (92.7) | 397 (92.5) |  |
| Single parent | 93 (5.0) | 16 (1.9) | 45 (7.3) | 32 (7.5) |  |
| Multiple adversities (≥2) |  |  |  |  | 170 (9.1) |
| No | 1585 (93.0) | 718 (94.6) | 517 (92.3) | 350 (90.9) |  |
| Yes | 119 (7.0) | 41 (5.4) | 43 (7.7) | 35 (9.1) |  |

### Associations between socioeconomic disadvantage, positive experiences, and poor CVH

Children with low or medium maternal education had a higher risk of poor CVH and fewer positive experiences than their high maternal education peers, after adjusting for baseline confounders (Table 2). Children who had fewer positive experiences had a higher risk (risk ratio (RR)=1.17; 95% CI=1.00, 1.36) of poor CVH than those who had two or more positive experiences, after adjusting for all confounders and maternal education. These findings confirm the hypothesized associations depicted in Figure 1.

**Table 2.** Generalized linear models examining the associations between maternal education, positive childhood experiences, and poor cardiovascular health, using imputed data (N=1874).

| Risk ratio (95% CI) |  |  |  |  |
| --- | --- | --- | --- | --- |
|  | Model 1:<br>Unadjusted | Model 2:<br>Adjusted for<br>baseline<br>confounders | Model 3:<br>Adjusted for<br>baseline and<br>intermediate<br>confounders | Model 4:<br>Adjusted for<br>baseline,<br>intermediate<br>confounders and<br>maternal<br>education |
| Association with poor cardiovascular health |  |  |  |  |
| Low maternal education<br>(ref=high) | 1.11 (0.92, 1.34) | 1.09 (0.91, 1.32) | - | - |
| Medium maternal education<br>(ref=high) | 1.11 (0.94, 1.30) | 1.11 (0.94, 1.31) | - | - |
| Fewer positive experiences<br>(ref=2 or more) | 1.18 (1.01, 1.37) | 1.17 (1.01, 1.36) | 1.17 (1.00, 1.36) | 1.17 (1.00, 1.36) |
| Association with fewer positive experiences |  |  |  |  |
| Low maternal education<br>(ref=high) | 1.23 (1.05, 1.45) | 1.19 (1.01, 1.41) | - | - |
| Medium maternal education<br>(ref=high) | 1.10 (0.95, 1.28) | 1.08 (0.93, 1.26) | - | - |

Baseline confounders controlled for were child's sex, child's ethnicity, child's disability, and neighborhood socioeconomic status. Intermediate confounders controlled for were gestational age in weeks, maternal age at childbirth, family composition, and childhood adversity. CI, confidence interval; -, not applicable.

### The extent to which intervening on positive experiences could reduce socioeconomic inequities in poor CVH

Using the interventional effects approach, we estimated an absolute difference of 4.9% (95% CI: −3.2%, 13.0%) and 5.6% (95% CI: −1.2%, 12.5%) in the prevalence of poor CVH in children with low and medium maternal education when compared with their high maternal education peers. If we were able to intervene to effectively increase the levels of positive experiences amongst children with low maternal education to be equivalent to their high maternal education peers, we could potentially reduce this absolute risk difference by 1% (95% CI: −0.8%, 1.5%). This translates to a relative reduction of 20.4% of socioeconomic inequities (Table 3). Similarly, hypothetical interventions that promote positive experiences amongst children with medium maternal education to be like their high maternal education peers, could potentially reduce the absolute risk difference by 0.5% (95% CI: −0.5%, 1.5%), leading to a relative reduction of 8.9% of socioeconomic inequities. After the hypothetical interventions, 3.9% and 5.1% absolute socioeconomic difference in CVH would remain respectively among children with low and medium maternal education.

**Table 3.**
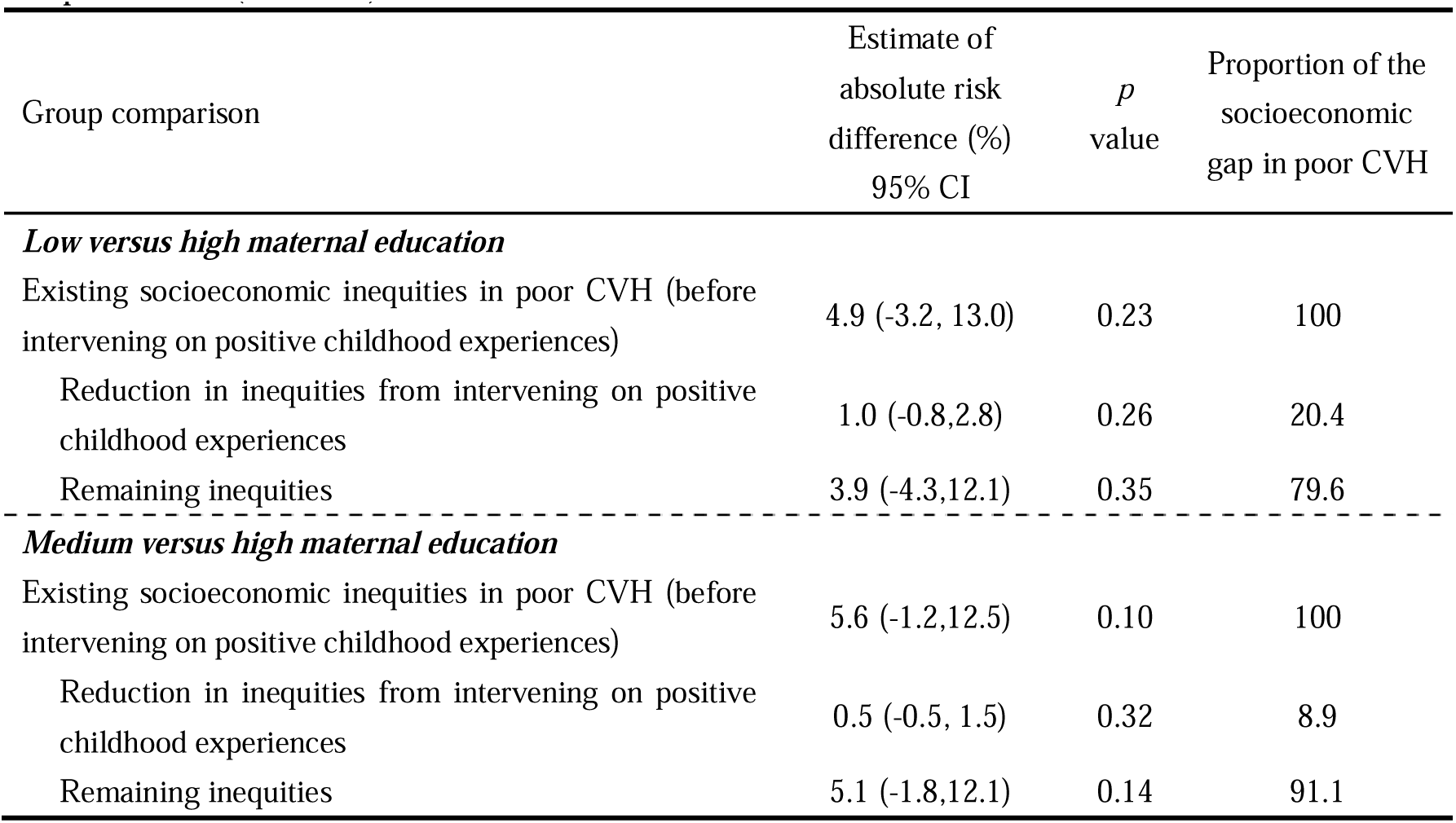
Results of evaluation of mediator interventions to close socioeconomic inequities in children’s poor cardiovascular health (CVH) using the interventional effects approach, using imputed data (N=1874).

| Group comparison | Estimate of<br>absolute risk<br>difference (%)<br>95% CI | *p*<br>value | Proportion of the<br>socioeconomic<br>gap in poor CVH |
| --- | --- | --- | --- |
| ***Low versus high maternal education*** |  |  |  |
| Existing socioeconomic inequities in poor CVH (before<br>intervening on positive childhood experiences) | 4.9 (-3.2, 13.0) | 0.23 | 100 |
| Reduction in inequities from intervening on positive<br>childhood experiences | 1.0 (-0.8,2.8) | 0.26 | 20.4 |
| Remaining inequities | 3.9 (-4.3,12.1) | 0.35 | 79.6 |
| ***Medium versus high maternal education*** |  |  |  |
| Existing socioeconomic inequities in poor CVH (before<br>intervening on positive childhood experiences) | 5.6 (-1.2,12.5) | 0.10 | 100 |
| Reduction in inequities from intervening on positive<br>childhood experiences | 0.5 (-0.5, 1.5) | 0.32 | 8.9 |
| Remaining inequities | 5.1 (-1.8,12.1) | 0.14 | 91.1 |

Results from the sensitivity analysis omitting the neighborhood socioeconomic status showed that promoting positive experiences in children with low and medium maternal education to be like their high maternal education peers could reduce the absolute risk difference by 1.2% and 0.6% respectively (see Supplementary file 6).

## DISCUSSION

Using prospective data from a national birth cohort, we estimated the potential reduction of socioeconomic inequities in children’s CVH if we could offer effective interventions to promote positive experiences among children with low or medium maternal education to be equivalent to their high maternal education peers. Our findings suggest a positive effect of positive experiences to reduce socioeconomic inequities in CVH, especially in children with low maternal education.

Consistent with previous findings,^21,39–41^ we found that children with more positive experiences had lower risk of poor CVH, after controlling childhood adversity and other confounders. We build on existing evidence by evaluating the potential benefit of a hypothetical intervention on positive experiences to reduce socioeconomic inequities in children’s CVH. Children with medium maternal education (5.6%) show a slightly greater absolute risk difference with their high maternal education peers than the low maternal education group (4.9%). However, promoting positive experiences appears to reduce a larger proportion of inequity among children with low maternal education. The greater benefit observed for children with low maternal education likely reflects the well-documented social gradient in CVH,^7^ where children from more disadvantaged families benefit more from interventions. Our estimates show that promoting positive experiences could reduce CVH inequities by up to 20.4%, highlighting the potential value of investing in positive experiences as a key intervention target. These reductions in childhood are likely to accumulate and translate to substantial social and health benefits in adulthood.^25^

Socioeconomic disadvantage can influence CVH through multiple interrelated pathways, including positive experiences as examined here, and other mediators such as childhood adversity, limited resources and healthcare access.^6,7^ Given this complexity, no single intervention will fully close the socioeconomic gap in children’s CVH. The residual inequities that persist after the hypothetical intervention on positive experiences suggest that a multi-faceted and stacked approach is needed to address both upstream and downstream factors of CVH.^42^

### Strengths and limitations

This study utilizes a national birth cohort that captured resourceful data on social, environmental, and physical measures longitudinally. We also used the target trial framework to provide clarity in the study design (e.g., eligibility criteria, treatment strategies). However, some limitations should be noted: (1) Selection bias: Although we conducted multiple imputation to reduce selection bias due to missing data in the sample, we were unable to account for the CheckPoint sampling weights in the analysis, meaning that some selection bias might remain and limit the generalisability of our findings; (2) Measurement bias: We used maternal education as a single measure of family socioeconomic position, which may underestimate the influence of socioeconomic disadvantage on the outcome. In addition, measurement errors may exist for parent-report or self-report measures, particularly with respect to the mediator and the outcome; and (3) Confounding bias: Despite adjusting for a range of potential confounders, residual or unmeasured confounding (e.g., racism, cultural norms, policy environment) remains a possibility.

### Implications for future research and practice

Our findings suggest that enhancing positive experiences has the potential to reduce socioeconomic inequities in children’s CVH. This study reinforces the importance of strengths-based approaches in epidemiolocal research that examine the positive health assets that allow populations to thrive, including in the face of adversity.^43,44^ We focused on a negative outcome in the present study; it is worthwhile to consider using a positive outcome in future to check whether results are consistent. It would be also interesting to explore the potential benefits of intervening on each type of positive experience to reduce socioeconomic inequities in each CVH component.^21^ Future work may also consider exploring the potential benefits in other countries and populations such as First Nations children.

The Australian Government’s *Early Years Strategy* highlights the importance of a strengths-based approach, leveraging positive resources to support children in reaching their optimal health.^15^ The hypothetical intervention in our study that would be capable of achieving an increase from “fewer than two” to “two or more” positive experiences remains undetermined (i.e., what the intervention is in practice and how to deliver it is not specified). Currently, there is increasing attention to programs targeting positive experiences to improve children’s health, such as the *Healthy Communities Study* in the US^45^ and the *Kids Building Future Healthy Mission* in Australia.^46^ It is likely to achieve the maximum impact by combining strategies that promote positive experiences with a multi-faceted and sustained approach that considers other fundamental drivers of CVH inequities.

## CONCLUSIONS

This study demonstrates that positive experiences partially mediate the relationship between socioeconomic disadvantage and poor CVH among Australian children. While promoting positive experiences has the potential to reduce socioeconomic inequities in CVH, addressing socioeconomic disadvantage itself and other socially distributed drivers of CVD remain imperative to achieve the maximum impact.

## Data Availability

All data produced in the present study are available upon reasonable request to the authors.

## Ethical approval

The LSAC (ID 13-04) and CheckPoint (ID 14-26) methodologies were approved by the Australian Institute of Family Studies Human Research Ethics Review Board, and the CheckPoint additionally by The Royal Children’s Hospital Melbourne Human Research Ethics Committee (33225D). This study was approved by the Royal Children’s Hospital Human Research Ethics Committee (ID 2019.170).

## Conflict of Interest Disclosures

The authors have indicated they have no conflicts of interest relevant to this article to disclose.

## Funding

This work is supported by the Victorian Government’s Operational Infrastructure Support Program. Dr Guo was supported by Murdoch Children’s Research Institute Population Health Theme Funding for 2023. Prof Burgner is supported by an NHMRC Investigator Grant (1175744). Prof Goldfeld is supported by an NHMRC Investigator Grant (2026263).

## Role of Funder

The funding sources had no role in the design and conduct of the study; collection, management, analysis, and interpretation of the data; preparation, review, or approval of the manuscript; and decision to submit the manuscript for publication.

## Abbreviations

AHA: American Heart Association
B-cohort: Birth cohort
CVD: Cardiovascular Disease
CVH: Cardiovascular Health
HOPE: Health Outcomes from Positive Experiences
LE8: Life’s Essential 8
LSAC: Longitudinal Study of Australian Children
RR: Risk Ratio

## Contributors’ Statement

Dr Shuaijun Guo conceptualized and designed the study, carried out data analyses, drafted the initial manuscript, and critically reviewed and revised the manuscript.

Dr Rushani Wijesuriya conceptualized and designed the study, carried out data analyses, and critically reviewed and revised the manuscript.

Drs Meredith O’Connor, Richard Liu, Profs David Burgner, and Sharon Goldfeld conceptualized and designed the study, and critically reviewed and revised the manuscript for important intellectual content.

Prof Naomi Priest conceptualized and designed the study, critically reviewed and revised the manuscript for important intellectual content and supervised Dr Guo.

All authors approved the final manuscript as submitted and agreed to be accountable for all aspects of the work.

## Notes

### Competing Interest Statement

The authors have declared no competing interest.

### Funding Statement

This study was funded by the Victorian Government's Operational Infrastructure Support Program and Murdoch Children's Research Institute Population Health Theme Funding for 2023.

### Author Declarations

The Royal Children's Hospital Human Research Ethics Committee gave ethical approval for this work (ID 2019.170).

