## Supplementary files for "Addressing Socioeconomic Inequities in Children’s Cardiovascular Health via Positive Experiences"

**Supplementary file 1.** Comparison of participants' characteristics between children attending CheckPoint and those not attending CheckPoint

Supplementary Table 1. Summary of participants' characteristics by whether children attended CheckPoint or not

| Participants' characteristics | Children attending CheckPoint | | |
| --- | --- | --- | --- |
|  | No (n=3233) | Yes (n=1874) | P value |
| ***Exposure*** |  |  |  |
| Maternal education | | |  |
| High | 851 (26.3) | 826 (44.1) | P<0.001 |
| Medium | 1198 (37.1) | 619 (33.0) |  |
| Low | 1181 (36.6) | 429 (22.9) |  |
| ***Confounders*** |  |  |  |
| Child's sex at birth |  |  |  |
| Female | 1580 (48.9) | 919 (49.0) | 0.908 |
| Male | 1653 (51.1) | 955 (51.0) |  |
| Child’s ethnicity at birth | |  |  |
| Anglo or European | 2544 (78.7) | 1616 (86.2) | P<0.001 |
| Minoritized ethnic group | 496 (15.3) | 221 (11.8) |  |
| Aboriginal and Torres Strait Islander | 193 (6.0) | 37 (2.0) |  |
| Child's disability status at birth |  |  |  |
| No | 3039 (94.0) | 1785 (95.3) | 0.060 |
| Yes | 194 (6.0) | 89 (4.7) |  |
| Neighborhood socioeconomic status at birth |  |  |  |
| Top 75% - Not disadvantaged | 2169 (67.1) | 1408 (75.1) | P<0.001 |
| Bottom 25% - Disadvantaged | 1064 (32.9) | 466 (24.9) |  |

**Supplementary file 2.** Rationale for the pathways specified in the conceptual model

Supplementary Table 2. Justifications for the inclusion of variables and pathways specified in the conceptual model, including justification of variable cut-offs.

| **Type of variable** | **Variable** | **Cut-off decision and citation** | **Relationship with exposure** | **Relationship with mediator and intermediate confounder** | **Relationship with outcome** |
| --- | --- | --- | --- | --- | --- |
| Exposure | Maternal education | Low or medium (disadvantaged) versus high (non-disadvantaged)^1^ | - - | - Children from disadvantaged families are likely to be exposed to fewer positive experiences.^2^ - Children with low maternal education are likely to be exposed to more adversities.^1^ - Low maternal education is associated with preterm birth.^3^ - Lower maternal education is associated with younger maternal age at childbirth.^4^ - Low maternal education is associated with family formation at an earlier age.^4^ | - Low maternal education is associated with worse children’s cardiovascular health.^5^ |
| Mediator | Positive childhood experiences | Two or more versus less than two^6,7^ | - - | - - | - Exposure to positive experiences is associated with better children’s cardiovascular health.^8^ |
| Outcome | Cardiovascular health | Poor versus good^9^ | - - | - - | - - |
| Baseline confounders | Child’s sex | Male versus female^10^ | - Child’s sex is a covariate that may influence maternal education.^1^ | - Child’s sex is a covariate that may influence positive experiences.^8^ - Child’s sex is a covariate that may influence gestational age.^8^ - Child’s sex is a covariate that may influence maternal age.^8^ - Child’s sex is a covariate that may influence family composition.^8^ | - Child’s sex is a covariate that may influence cardiovascular health.^8^ |
|  | Child’s ethnicity | Anglo or European versus minoritized ethnic group versus Indigenous^8^ | - Child’s ethnicity is a covariate that may influence maternal education.^1^ | - Child’s ethnicity is a covariate that may influence positive experiences.^8^ - Child’s ethnicity is a covariate that may influence gestational age.^8^ - Child’s ethnicity is a covariate that may influence maternal age.^8^ - Child’s ethnicity is a covariate that may influence family composition.^8^ | - Child’s ethnicity is a covariate that may influence cardiovascular health.^8^ |
|  | Child’s disability status | Yes versus no^10^ | - Child’s disability is a covariate that may influence maternal education.^11^ | - Child’s disability is a covariate that may influence positive experiences.^8^ - Child’s disability is a covariate that may influence gestational age ^8^ - Child’s disability is a covariate that may influence maternal age.^8^ - Child’s disability is a covariate that may influence family composition.^8^ | - Child’s disability is a covariate that may influence cardiovascular health.^8^ |
|  | Neighbourhood socioeconomic status | Top 75% - not disadvantaged versus bottom 25% - disadvantaged^10^ | - Neighbourhood socioeconomic status is a covariate that may influence maternal education.^12^ | - Neighbourhood socioeconomic status is a covariate that may influence positive experiences.^8^ - Neighbourhood socioeconomic status is a covariate that may influence gestational age.^8^ - Neighbourhood socioeconomic status is a covariate that may influence maternal age.^8^ - Neighbourhood socioeconomic status is a covariate that may influence family composition.^8^ | - Neighbourhood socioeconomic status is a covariate that may influence cardiovascular health.^8^ |
| Intermediate confounder | Gestational age in weeks | <37 weeks versus ≥37 weeks^13^ | - - | - Gestational age in weeks is a covariate that may influence positive experiences.^8^ | - Preterm birth is associated with poor cardiovascular health.^14^ |
|  | Maternal age at childbirth | <27 years versus ≥27 years^10^ | - - | - Maternal age at childbirth is a covariate that may influence positive experiences.^8^ | - Advanced maternal age is associated with poor cardiovascular health.^15^ |
|  | Family composition | Yes versus no^10^ | - - | - Family composition is a covariate that may influence positive experiences.^8^ | - Single parenthood is associated with poor cardiovascular health.^16^ |
|  | Adverse childhood experiences | Two or more versus less than two^7^ | - - | - Childhood adversity is a covariate that may influence positive experiences.^8^ | - Exposure to multiple adversities is associated with poor cardiovascular health.^8^ |

**Supplementary file 3.** Measurement of positive and adverse childhood experiences

Supplementary Table 3. Measures used to define positive and adverse experiences

|  | **Measurement and example item** | **Age assessed** |
| --- | --- | --- |
| ***Positive experiences (17 indicators)*** | |  |
| Warm parenting | Six items derived from the original 9-item Childrearing Questionnaire (Paterson & Sanson, 1999; Sanson, 1995), reported by P1 and P2. E.g. “How often do you express affection by hugging, kissing and holding this child?” Mean of six items at each time point was derived for each parent, and the top 25% was coded as warm parenting,^17^ to identify relatively higher levels of these behaviours. No parent reporting warm parenting=0; P1 and/or P2 reporting high levels=1. | 2-3 years, 4-5 years, 6-7 years, 8-9 years, 10-11 years |
| High co-parenting alliance | Three items adapted from the Quality of Co-parental Interaction Scale (Ahrons, 1981); reported by P1 and P2. E.g. “How often is your partner a resource or support to you in raising your child?” Mean of three items at each time point was derived for each parent, and the top 25% was coded as high co-parenting alliance. No parent reporting high co-parenting alliance =0; P1 and/or P2 reporting high levels =1. Single parent was coded as 0. | 2-3 years, 4-5 years, 6-7 years, 8-9 years, 10-11 years |
| Consistent parenting | Five items derived from the National Longitudinal Survey of Children and Youth (NLSCY): Cycle 4 (Survey Instruments, 2000-2001, Parent Questionnaire), reported by P1 and P2. E.g. “When you give this child an instruction or make a request to do something, how often do you make sure that he/she does it?” Mean of five items at each time point was derived for each parent, and the top 25% was coded as high consistent parenting. No parent reporting high consistent parenting=0; P1 and/or P2 reporting high consistent parenting =1. | 4-5 years, 6-7 years, 8-9 years, 10-11 years |
| High family relationship quality | Wave 3 to 6: Six items derived from the Relationship Assessment Scale [RAS] (Hendrick, 1988); Wave 2: Two items derived from the Early Childhood Longitudinal Study-Birth Cohort (US Department of Education), reported by P1 and P2. E.g. “How well does your partner meet your needs?” High family relationship quality at each time point was derived for each parent, and the top 25% was coded as high. No parent reporting high family relationship quality=0; P1 and/or P2 reporting high levels =1. Single parent was coded as 0. | 2-3 years, 4-5 years, 6-7 years, 8-9 years, 10-11 years |
| High parental support for raising children | Wave 2 to 6: Five LSAC designed items, reported by P1 and P2. E.g. “How often do your parents support you in raising your child?” Mean of five items at Wave 2 to 6 was derived for each parent, and the top 25% was coded as high parental support for raising children. No parent reporting high parental support for raising children=0; P1 and/or P2 reporting high parental support for raising children =1. | 2-3 years, 4-5 years, 6-7 years, 8-9 years, 10-11 years |
| High social support for study child (SC) | Seven items adapted from the British Cohort Study (Centre for Longitudinal Studies, 2004), reported by SC. E.g. “If you had a problem, who would you talk to about it? Mum” Mean of seven items at 10-11 years was derived for study child, and the top 25% was coded as high social support for study child. Bottom 75%=0; top 25%=1. | 10-11 years |
| High quality parent relationship with SC | Eight items derived from the Trust & communication scale which was drawn from the People in My Life measure (PIML); Ridenour, Greenberg & Cook (2006), reported by SC. E.g. “My parents accept me as I am.” Mean of eight items at 10-11 years was derived for study child, and the top 25% was coded as positive parent relationship for study child. Bottom 75%=0; top 25%=1. | 10-11 years |
| High teacher/carer relationship with SC | Three or four items derived from the Student-Teacher Relationship Scale (STRS) (Pianta, 1991), reported by Teacher/carer. E.g. “I share an affectionate, warm relationship with this child” Mean of three items at 4-9 years and mean of four items at 10-11 years were derived for study child, and the top 25% was coded as positive teacher/carer relationship for study child. Bottom 75%=0; top 25%=1. | 4-5 years, 6-7 years, 8-9 years, 10-11 years |
| High peer relationship with SC | Eight items derived from the Marsh Self Description Questionnaire II (Marsh, 1990), reported by SC. E.g. “I have many friends.” Mean of eight items at Wave 5 to 6 were derived for study child, and the top 25% was coded as high peer quality. Bottom 75%=0; top 25%=1. | 8-9 years, 10-11 years |
| High neighbourhood liveability | Three or four items derived from AIFS Families, Social Capital and Citizenship survey (Stone & Hughes, 2002) and the NSW ‘Communities 4 Kids’ initiative / WA Child Health Survey, reported by P1. E.g. “There are good parks, playgrounds and play spaces in this neighbourhood.” Mean of items at each time point was derived for P1, and the top 25% was coded as high neighbourhood liveability. Bottom 75%=0; top 25%=1. | 2-3 years, 4-5 years, 6-7 years, 8-9 years, 10-11 years |
| High accessibility of neighbourhood facilities | Three items designed by LSAC, based on work from the WA Child Health Survey, AIFS Families, Social Capital and Citizenship survey, and the NSW ‘Communities 4 Kids’ initiative / WA Child Health Survey, reported by P1. E.g. “There is access to close, affordable, regular public transport in this neighbourhood.” Mean of three items at each time point was derived for P1, and the top 25% was coded as supportive neighbourhood facilities. Bottom 75%=0; top 25%=1. | 2-3 years, 4-5 years, 6-7 years, 8-9 years, 10-11 years |
| High neighbourhood social capital | Wave 2 to 6: Two items derived from the WA Child Health Survey and the National Longitudinal Survey of Children and Youth (NLSCY): Cycle 1, reported by P1. E.g. “It is safe for children to play outside during the day.” Mean of two items at Wave 2 to 6 was derived for P1, and the top 25% was coded as high neighbourhood social capital. Bottom 75%=0; top 25%=1. | 2-3 years, 4-5 years, 6-7 years, 8-9 years, 10-11 years |
| Positive home education environment for SC | Wave 2 to 5: Two or seven items derived from a range of longitudinal study surveys including the Early Childhood Longitudinal Study – Birth Cohort K Base Year instruments, Head Start Family and Child Experiences Survey, the National Household Education Survey and the Longitudinal Literacy and Numeracy Study; Wave 6: Two items adapted from the 'Millennium Cohort Study (MCS4); Centre for Longitudinal Studies (2008), reported by P1. E.g. “In the past week, on how many days have you or an adult in your family, read to child from a book?” Mean of items at each time point was derived for P1, and the top 25% was coded as positive home learning environment. Bottom 75%=0; top 25%=1. | 2-3 years, 4-5 years, 6-7 years, 8-9 years, 10-11 years |
| High frequency of contact with family and friends for SC | Five or six LSAC designed items, based on frequency of contact questions contained in the National Statistics Omnibus Survey 2002 (Non-resident parental contact module) and the Early Childhood Longitudinal Study, Birth Cohort, ECLS-B, [US Department of Education] (Non-resident Father Questionnaire), reported by P1. E.g. “How often does the study child see or spend time with your neighbors?” Mean of items at each time point was derived for P1, and the top 25% was coded as high frequency of contact with family and friends for study child. Bottom 75%=0; top 25%=1. | 2-3 years, 4-5 years, 6-7 years, 8-9 years, 10-11 years |
| High number of activities outside the home for SC | Wave 2 to 6: Five items derived from the National Longitudinal Survey of Children and Youth (NLSCY): Cycle 3 (Survey Instruments, 1998-1999, Parent Questionnaire); reported by P1. E.g. “In the past month, has child visited a library with you or another family member?” Sum of items at each time point was derived for P1, and the top 25% was coded as positive learning environment outside the home. Bottom75%=0; top 25%=1. | 2-3 years, 4-5 years, 6-7 years, 8-9 years, 10-11 years |
| High care/school enjoyment | Wave 3: Four items derived from Leiden Inventory for the Child’s Well-being in Day Care (LICW-D); Wave 4 to 6: Nine or twelve items derived from School Liking & Avoidance Scale, adapted from the School Sentiment Inventory (Ladd & Price, 1987), reported by Teacher/carer. E.g. “How often does study child appear to look forward to going to school?” Mean of items at each time point was derived for care enjoyment, and the top 25% was coded as high enjoyment. Bottom 75%=0; top 25%=1. | 4-5 years, 6-7 years, 8-9 years, 10-11 years |
| Strong sense of neighbourhood belonging | Four items adapted the Project on Human Development, Chicago Neighborhoods: Community Survey, 1994-1995, Instrument for ICPSR 2766, AIFS Families, Social Capital and Citizenship survey (Stone & Hughes, 2002) and the NSW ‘Communities 4 Kids’ initiative / WA Child Health Survey; reported by P1. E.g. “Most people in your neighbourhood can be trusted.” Mean of items at each time point was derived for P1, and the top 25% was coded as high sense of belonging. Bottom75%=0; top 25%=1. | 2-3 years, 4-5 years, 6-7 years, 8-9 years, 10-11 years |
| ***Adverse experiences (8 indicators)*** | |  |
| Parent legal problems | A single item from the stressful life events scale adapted from Brugha and Cragg (1990), reported by P1: “In the last year, have any of the following happened to you? You had problems with the police and a court appearance.” No=0; Yes=1. | 0-1 year |
| Interparental violence | A single item from an adapted version of the Quality of Co-parental Interaction Scale (Ahrons, 1981), reported by P1 and P2: “How often do you have arguments with your partner that end up with people pushing, hitting, kicking or shoving?” ‘Never’=0, ‘Rarely’ to ‘Always’=1. Single parent was coded as 0. | 0-1 year |
| Household member mental illness | The six-item K-6 Depression Scale reported by P1 and P2; provides a brief measure of psychological distress. Items asked in reference to the past 30 days, e.g., “During the past 30 days, about how often did you feel hopeless?” Score over 13 (mental disorder very likely) categorized as high psychological distress. Neither parent high distress=0; P1 and/or P2 high distress=1. | 0-1 year |
| Household member substance abuse | As for Parent legal problems, reported by P1: “In the last year, have any of the following happened to you? Someone in your household had an alcohol or drug problem” No=0; Yes=1. | 0-1 year |
| Harsh parenting | Harsh parenting measured using three adapted items from the Early Childhood Longitudinal Study of Children, Birth Cohort and the National Longitudinal Survey of Children and Youth 1998-1999, reported by P1 and P2. E.g. “How often do you tell this child that he/she is bad or not as good as others?” Mean of items at each time point was derived for each parent, and the top 5% within each wave was coded as harsh parenting, to identify relatively higher levels of these behaviours. Neither parent reporting harsh parenting=0; P1 and/or P2 reporting high levels=1. | 0-1 year |
| Parental separation/divorce | As for Parent legal problems, reported by P1: “In the last year, have any of the following happened to you? You had a separation due to relationship or marital difficulties.” No=0; Yes=1. | 0-1 year |
| Unsafe neighbourhood | A single item informed by the WA Child Health Survey, AIFS Families, Social Capital and Citizenship survey and the NSW ‘Communities 4 Kids’ initiative / WA Child Health Survey, reported by P1: “How strongly do you agree or disagree with these statements about your neighbourhood? This is a safe neighbourhood.” ‘Strongly agree’ or ‘agree’ = 0; ‘disagree’ or ‘strongly disagree’ = 1. | 0-1 year |
| Family member death | As for Parent legal problems, item reflecting death of a parent, partner or child, reported by P1: “In the last year, have any of the following happened to you? Your parent, partner or child died.” No=0; Yes=1. | 0-1 year |

**Supplementary file 4.** Measurement of children’s cardiovascular health

Supplementary Table 4.1 Scoring approach for quantifying children’s cardiovascular health in LSAC, as per the American Heart Association’s Life’s Essential 8 score^9^

| **Domain** | **CVH Metric** | **Method of measurement** | **Data source** | **Quantification of CVH Metric for children** |
| --- | --- | --- | --- | --- |
| Health behaviours | Diet | Self-reported daily intake of diet pattern, using the Modified National Secondary Students’ Diet and Activity (NaSSDA) questionnaire | CheckPoint (2015) | Metric: See Supplementary Table 2.2 for details  Scoring (population):   - ≥ 95^th^ percentile (top/ideal diet; 100 points) - 75^th^-94^th^ percentile (80 points) - 50^th^-74^th^ percentile (50 points) - 25^th^-49^th^ percentile (25 points) - 1^st^-24^th^ percentile (bottom/least ideal diet; 0 point) |
|  | Physical activity | Self-reported minutes of moderate or vigorous physical activity per week, using the Multimedia Activity Recall for Children and Adults (MARCA) questionnaire | CheckPoint (2015) | Metric: Minutes of moderate- (or greater) intensity activity per week  Scoring:   - ≥ 420 minutes (100 points) - 360-419 minutes (90 points) - 300-359 minutes (80 points) - 240-299 minutes (60 points) - 120-239 minutes (40 points) - 1-119 minutes (20 points) - 0 minute (0 point) |
|  | Nicotine exposure | Self-reported use of cigarettes and parent-reported use of cigarettes, using questions from the Australian School Students Alcohol and Drug Survey, the Western Australian Aboriginal Child Health Survey - Carer's Questionnaire, and Kemper self-administered questionnaire for Family Psychosocial Screening | LSAC Wave 1-7 (2004-2016) | Metric: Cigarettes smoking or second-hand smoke exposure  Scoring:   - Never tried smoking (100 points) - Tried smoking but just a few puffs (50 points) - Tried smoking but fewer than 10 cigarettes (25 points) - Tried smoking 10 to 100 cigarettes (0 points)   Subtract 20 points (unless score is 0) for living with active indoor smoker in home |
|  | Sleep health | Self-reported minutes of sleep average time per night, using the Multimedia Activity Recall for Children and Adults (MARCA) questionnaire | CheckPoint (2015) | Metric: Average hours of sleep per night  Scoring:   - Age-appropriate optimal range: 9-12 hours (100 points) - < 1 h above optimal range (90 points) - < 1 h below optimal range (70 points) - 1-< 2 h below or ≥ 1 h above optimal range (40 points) - 2-< 3 h below optimal range (20 points) - ≥ 3 h below optimal range (0 point) |
| Health factors | Body mass index (BMI) | Body weight (kg) divided by height squared (m^2^), using Stadiometer (height) & InBody230 BIA scale (weight) & the US Centers for Disease Control and Prevention (CDC) reference values for children | CheckPoint (2015) | Metric: BMI percentiles for age and sex  Scoring:   - 5^th^-<85^th^ percentile (100 points) - 85^th^-95^th^ percentile (70 points) - 95^th^ percentile-<120% of the 95^th^ percentile (30 points) - 120% of the 95^th^ percentile -<140% of the 95^th^ percentile (15 points) - ≥ 140% of the 95^th^ percentile (0 point) |
|  | Blood lipids^*^ | Serum total and high-density lipoprotein (HDL)-cholesterol with calculation of non-HDL-cholesterol, using the Nuclear Magnetic Resonance (NMR) metabolomics platform | CheckPoint (2015) | Metric: Non–HDL cholesterol (mg/dL)  Scoring:   - < 100 mg/dL (100 points) - 100-119 mg/dL (60 points) - 120-144 mg/dL (40 points) - 145-189 mg/dL (20 points) - ≥ 190 mg/dL (0 points) |
|  | Blood glucose^*^ | Non-fasting average blood glucose, using the NMR metabolomics platform. HbA1c to average blood glucose (mg/dL) conversion: (HbA1c × 35.6) - 77.3 = Average blood glucose | CheckPoint (2015) | Metric: Average blood glucose (mg/dL)  Scoring:   - No history of diabetes and average blood glucose < 100 mg/dL (100 points) - No diabetes and average blood glucose 100-125 mg/dL (prediabetes; 60 points) - Diabetes with average blood glucose 126-172 mg/dL (40 points) - Diabetes with average blood glucose 173-204 mg/dL (30 points) - Diabetes with average blood glucose 205-240 mg/dL (20 points) - Diabetes with average blood glucose 241-275 mg/dL (10 points) - Diabetes with average blood glucose ≥ 276 mg/dL (0 point) |
|  | Blood pressure (BP) | Appropriately measured systolic and diastolic blood pressure, using the Sphygmocor XCEL & *The Fourth Report* reference values for children | CheckPoint (2015) | Metric: Systolic and diastolic BP (mm Hg) percentiles for age  Scoring:   - Optimal: <90^th^ percentile (100 points) - Elevated: ≥90^th^-<95^th^ percentile or ≥120/80 mm Hg to <95^th^ percentile, whichever is lower (75 points) - Stage 1 hypertension: ≥95^th^-<95^th^ percentile+12 mm Hg, or 130/80 to 139/89 mm Hg, whichever is lower (50 points) - Stage 2 hypertension: ≥95^th^ percentile+12 mm Hg or ≥140/90 mm Hg, whichever is lower (25 points) - Systolic BP ≥160 or ≥95^th^ percentile+30 mm Hg systolic BP, whichever is lower; and/or diastolic BP≥100 or ≥95^th^ percentile+20 mm Hg diastolic BP (0 point) |

* Multiply mg/dL values to obtain mmol/L, by 0.02586 for cholesterol, and by 0.0555 for glucose.

Supplementary Table 4.2 Scoring criteria for diet score

| Component | Measurement item | Scoring criteria* |
| --- | --- | --- |
| Fruit | How many serves of FRUIT you USUALLY eat each day? Do not include fruit juice. Include all fresh, dried, frozen, and tinned fruit. | ≥2 servings of fruit per day (20 points) |
| Vegetables | How many serves of VEGETABLES you USUALLY eat each day? Do not include any potatoes, hot chips or fried potato. Include all fresh, dried, frozen and tinned vegetables. | ≥5 serves of vegetables per day (20 points) |
| Whole grains | 1) How many slices of BREAD you USUALLY eat each day? 2) What type of BREAD do you USUALLY eat? If you usually eat more than one type of bread, select the one you eat most often. | ≥2 slices of wholegrain bread per day (20 points) |
| Red and processed meats | How often do you eat RED MEAT such as beef or lamb? Include all steaks, chops, roasts, mince, stir fries and casseroles? (Do not include pork or chicken). | Less than once per week (20 points) |
| Sweetened beverages | 1) How much FRUIT JUICE do you USUALLY drink? 2) How much SOFT DRINKS like Coke, lemonade, CORDIALS or SPORTS DRINKS like Gatorade do you USUALLY drink? Do not include diet soft drinks. 3) How much ENERGY DRINKS like Redbull, V, do you USUALLY drink? | <4 cups per week (20 points) |

* Scoring criteria were adapted to available National Secondary Students Diet and Activity Questionnaire data, Australian Dietary Guideline of healthy eating for children, and American Heart Association recommendations.

**Supplementary file 5.** Additional details about the outcome and mediator models used for estimating the interventional effects of hypothetical interventions using extended g-computation.

In the primary analysis (adjusted for baseline confounders child’s sex at birth, ethnicity, disability status, and neighborhood socioeconomic status and intermediate confounders gestational age, maternal age at childbirth, family composition and multiple childhood adversities), the outcome model included the exposure (poor cardiovascular health), mediator (positive childhood experiences), intermediate confounders, baseline confounders, and two-way interactions between exposure and mediator and, mediator and intermediate confounders. The model for the mediator included exposure, baseline confounders, intermediate confounder, while the model for intermediate confounders included the exposure, baseline confounders.

In the sensitivity analysis , the outcome, and mediator models remained the same, except excluding the baseline confounder neighborhood socioeconomic status .The joint distribution of the intermediate confounders was modelled by decomposing the joint distribution into sequential conditional distributions assuming a non-causal order as follows:

- Regression for $L_{4}$on exposure, baseline confounders
- Regression for $L_{3}$on exposure, baseline confounders, $L_{4}$
- Regression for $L_{2}$on exposure, baseline confounders, $L_{4},L_{3}$
- Regression for $L_{1}$on exposure, baseline confounders,$, L_{4},L_{3},L_{2}$

Where $L_{1}$: gestational age, $L_{2}$: maternal age at childbirth, $L_{3}:$family composition, and $L_{4} :$multiple childhood adversities

**Supplementary file 6:** Results of evaluation of mediator interventions to close socioeconomic inequities in children’s poor cardiovascular health using the interventional effects approach, using imputed data (N=1874), sensitivity analysis omitting neighborhood socioeconomic status from the baseline confounder set.

| Group comparison | | Estimate of absolute risk difference (%)  95% CI | *p*  value | Proportion of the socioeconomic gap in poor CVH |
| --- | --- | --- | --- | --- |
| ***Low versus high maternal education*** | |  |  |  |
| Existing socioeconomic inequities in poor CVH (before intervening on positive childhood experiences) | | 5.5 (-2.4, 13.4) | 0.17 | 100 |
|  | Reduction in inequities from intervening on positive childhood experiences | 1.2 (-0.8,3.2) | 0.25 | 21.8 |
|  | Remaining inequities | 4.3 (-3.7, 12.3) | 0.29 | 78.2 |
| ***Medium versus high maternal education*** | |  |  |  |
| Existing socioeconomic inequities in poor CVH (before intervening on positive childhood experiences) | | 6.0 (-0.7,12.7) | 0.08 | 100 |
|  | Reduction in inequities from intervening on positive childhood experiences | 0.6 (-0.4, 1.7) | 0.26 | 10 |
|  | Remaining inequities | 5.4 (-1.4, 12.2) | 0.12 | 90 |
